# Loss of chromosome Y in regulatory T cells

**DOI:** 10.1101/2023.06.17.23291316

**Authors:** Jonas Mattisson, Jonatan Halvardson, Hanna Davies, Bożena Bruhn-Olszewska, Paweł Olszewski, Marcus Danielsson, Josefin Bjurling, Amanda Lindberg, Ammar Zaghlool, Edyta Rychlicka-Buniowska, Jan P. Dumanski, Lars A. Forsberg

**Author notes:** Authors with equal contribution (Jonas Mattisson, Jonatan Halvardson).

## Abstract

Mosaic loss of chromosome Y (LOY) in leukocytes is the most prevalent somatic aneuploidy in aging humans. Men with LOY have increased risks of all-cause mortality and the major causes of death, including many forms of cancer. It has been suggested that the association between LOY and disease risk depends on what type of leukocyte is affected with Y loss, with prostate cancer patients showing higher levels of LOY in CD4+ T lymphocytes. In previous studies, Y loss has however been observed at relatively low levels in this cell type. This motivated us to investigate whether specific subsets of CD4+ T lymphocytes are particularly affected by LOY. Studying publicly available, T lymphocyte enriched, single-cell RNA sequencing datasets from cancer patients, we found that regulatory T cells (Tregs) had significantly more LOY than any other T lymphocytes studied. To validate these findings, we generated a single-cell RNA sequencing dataset comprised of 23 PBMC samples and 32 CD4+ T lymphocytes enriched samples. We observed an increased ratio of Tregs with LOY compared with other CD4+ cells, supported by developmental trajectory analysis of CD4+ T lymphocytes culminating in the Treg subtype. Furthermore, we identify dysregulation of 465 genes in Tregs with Y loss, many involved in the immunosuppressive functions and development of Tregs. Considering that Tregs plays a critical role in the process of immunosuppression; this enrichment for Tregs with LOY might contribute to the increased risk for cancer observed among men with Y loss in leukocytes.

## INTRODUCTION

Loss of chromosome Y from hematological progenitors results in mosaic loss of chromosome Y (LOY) in circulating leukocytes, representing the most prevalent somatic mutation in general populations^1-3^. Affected men have an increased risk for mortality and morbidity^4,5^, including all major causes of death such as cardiovascular disease^5-7^, cancer^2,4,8^, Alzheimer’s disease^9^. The link between Y loss in blood and disease in other organs is currently being explored, with evidence emerging for direct causality related to the LOY condition. It has for instance been shown that LOY: I) affects the distribution of blood cell types^10,11^, II) leads to dysregulation of almost 500 autosomal genes in a cell type dependent manner^12^, III) reduces the abundance of immunoprotein CD99 on the surface of cells, a protein crucial for regulating the permeability of blood vessels^13^, IV) induce fibrosis of internal organs and organ dysfunction in murine models^7^, V) is linked to an expansion of low-density neutrophils during COVID-19 infection^14^. Furthermore, we have previously shown that the impact of LOY on disease risk depends on which leukocyte subset is affected by Y loss^12^. Specifically patients with Alzheimer’s disease had more LOY in NK cells, while patients with prostate cancer displayed higher levels of LOY in granulocytes as well as CD4+ T lymphocytes^12^. This however results in an enigma: Y loss in CD4+ T lymphocytes is associated with increased risk of cancer, yet LOY occurs at relatively low levels in CD4+ T lymphocytes compared with other leukocytes^3,12,13^.

Regulatory T cells (Tregs) are a highly specialized subtype of CD4+ T lymphocytes, constituting less than 10% of circulating CD4+ cells^15,16^. Hence, the total population of peripheral blood mononuclear cells (PBMC) contains an even smaller fraction of Tregs, which can be difficult to discern from other CD4+ T lymphocytes in single-cell experiments^15,16^. Yet, Tregs play a critical role in the regulation of immune functions by performing immunosuppression, a balancing act as not to suppress a necessary immune response^17^. Thus, Treg dysregulation has been linked to processes such as long-term inflammation, tissue damage and increased fibrosis^18^. Tregs also contribute to tumour development by directly inhibiting the immune response of surrounding effector cells^19,20^. Considering the major impact of this rare CD4+ subset, we sought to investigate how the heterogeneity of CD4+ T lymphocytes is affected by LOY. Here, we leverage the power of single-cell RNA sequencing (scRNAseq) in multiple datasets, as well as T lymphocyte enrichment, to study how Y loss influences the developmental trajectory, distribution and gene expression of this rare leukocyte population.

## RESULTS

### Distribution of LOY in T lymphocyte subsets

To investigate the distribution of LOY cells in CD4+ T lymphocytes we collected three public scRNAseq datasets containing fluorescence-activated cell sorted (FACS) T lymphocytes from cancer patients, with samples taken from tumour, healthy tissue and blood^21-23^. After standard pre-processing and clustering 21,709 cells remained, out of which 8,647 originated from male donors. Known marker genes were used to assign cell types (Supplementary Figure 1A) to each cluster (Supplementary Figure 1B) based on all 21,709 cells. The abundance of reads from the male specific region of the Y chromosome (MSY) was thereafter used to assign LOY status to each of the 8,647 cells from male donors. Due to the nature of the dataset, some cells from female donors also had MSY reads. Thus, the expression from MSY in female cells was considered as technical noise and used as a threshold when estimating the LOY status of single-cells from men. For each cell type the percentage of LOY-cells was calculated, showing LOY at marginal levels for the majority of the T lymphocytes. Strikingly, Tregs had a significant elevation of LOY with a close to three times higher occurrence of Y loss compared to other types of T lymphocytes (Wilcoxon signed-rank test: p = 0.00061, Figure 1A).

**Figure 1:**
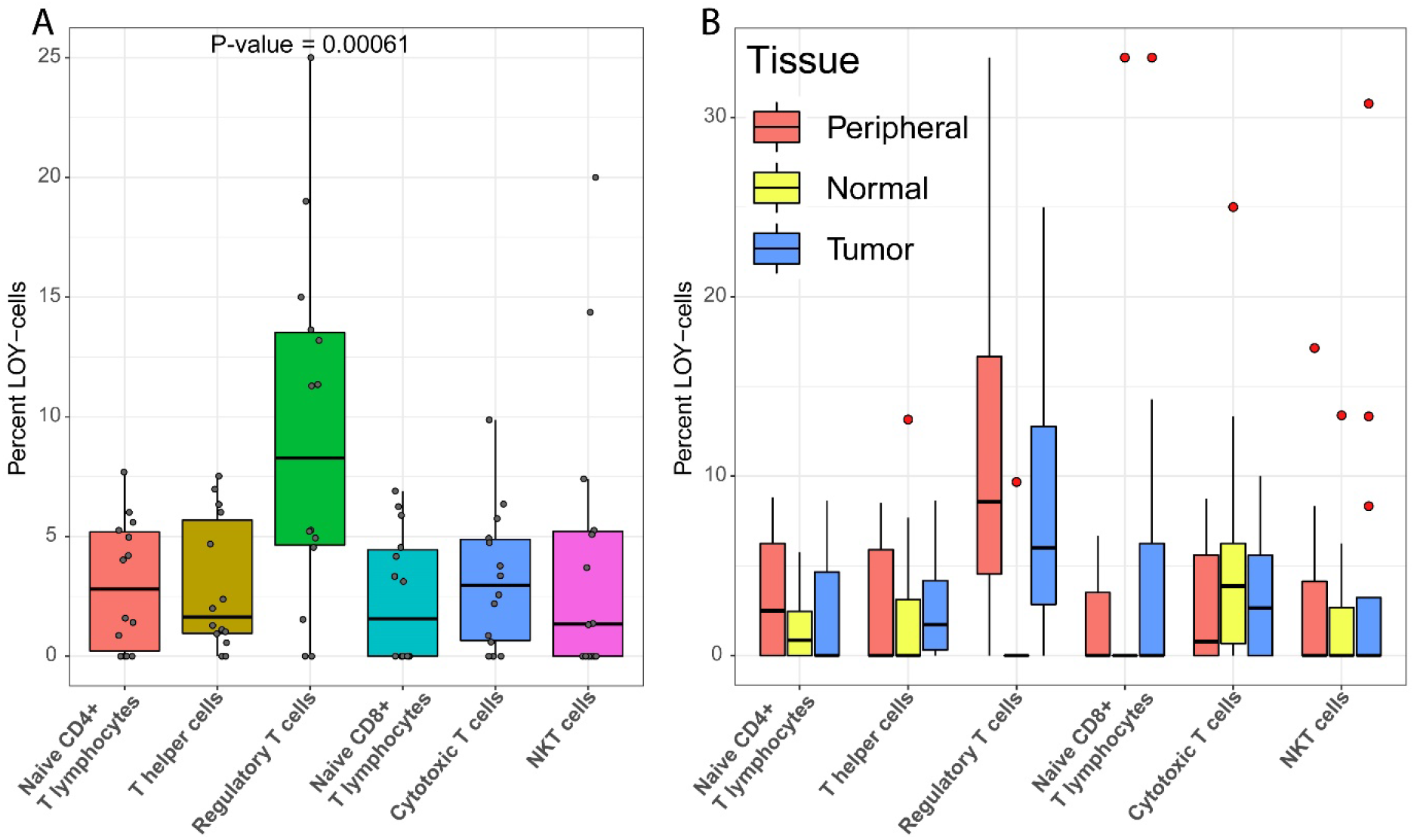
**A**) Boxplot showing the overall percentage of LOY in different T lymphocyte subsets, in each studied subject of the public datasets. Each dot corresponds to the LOY frequency of one patient, color-coded for each cell type. **B**) Similar boxplot as in *A*, stratified by tissue of origin for T lymphocytes as indicated. Peripheral, Normal and Tumour indicates peripheral blood, normal tissue adjacent to tumour and tumour tissue, respectively. Note that the dots here marks outliers, and has therefore been coloured red to differentiate them from the dots in *A*.

Given the observation that Tregs are the main carriers of LOY in the public datasets, we sought to investigate if Y loss might influence the distribution of T lymphocyte subtypes. First we used linear regression to test whether the fraction of Tregs (relative to the total number of CD4+ T lymphocytes) in each subject was associated with the percentage of LOY in Tregs. This analysis showed that subjects with higher level of LOY in Tregs had a larger fraction of Tregs compared with other subjects (Coeff = 2.622, p = 0.033965). Next, we evaluated this result in a model adjusting for relevant confounding factors such as dataset and individual. In support of the unadjusted model, a negative binomial model showed that the number of cells within each cell type was significantly associated with the level of LOY (GLM: LR = 6.591, Df = 1, p = 0.01025). Other significant confounders included cell type, as well as the interaction between Y loss and cell type (Supplementary Table 1). Finally, we also investigated how LOY varied between the sampled tissues (Figure 1B). While LOY-cells were identified in all cell types and tissues, Tregs and CD8+ T lymphocytes with LOY were principally absent from normal tissue. To test the association between Y loss and sampled tissue, a quasibinomial model was used. This test confirmed that the level of LOY varied between tissues (GLM: LR = 8.426, Df = 2, p = 0.014804), as well as cell type, cancer type and the interactions between cell type, cancer type and tissue (Supplementary Table 2). Together, these results suggest that LOY in T lymphocytes might influence the distribution of T lymphocyte subsets. To further investigate a potential effect from Y loss on T lymphocyte development, we generated a larger scRNAseq dataset from an independent cohort as described below.

### Validation of LOY distribution in an independent cohort

We performed scRNAseq on peripheral blood mononuclear cells (PBMCs), from freshly collected blood samples, from 55 males in the EpiHealth, UCAN and the UAD cohort (see methods). Enrichment for CD4+ T lymphocytes was performed in 32 of these (from the EpiHealth and UCAN cohorts) prior to sequencing to achieve a larger fraction of Tregs in the studied cell population. After standard QC of the scRNAseq data, the final dataset consisted of all 55 samples and 213,619 single-cells, grouped into 27 clusters based on RNA expression profiles (Supplementary Figure 2A). The major cell type in each cluster was identified combining two different machine learning approaches, CIPR and singleCellNet. Both classification tools were trained on publicly available datasets (see methods) and used to predict the most likely cell type of each cluster. The final cell type classification was generated using the resulting predictions combined with manual curation based on the expression of known marker genes (Supplementary Figure 2B). Next, LOY status was estimated for each cell separately, classifying cells without any MSY reads as LOY-cells. This method considers all reads within transcripts from MSY genes, unlike our previously published method, which only considered spliced RNA^12^. By applying a published benchmarking method for LOY scoring^24^ that considers the overall MSY gene expression in each cluster, we found that our new method of LOY calling had considerably improved accuracy (Supplementary Figure 3 & 4).

The frequency of LOY was thereafter characterized in the different types of studied leukocytes (Figure 2A). In line with previous results, NK cells and monocytes exhibited the highest percentage of Y loss in individual samples^3,12,13^. However, Tregs had the highest median LOY value (17.91%) of all cell types in the validation dataset (Figure 2A), replicating the observation from the public dataset (Figure 1A). Furthermore, Tregs showed a significantly higher LOY fraction than other T cells (Wilcoxon signed-rank test: p = 1.139e-10, Figure 2B) and was the only CD4+ subset to harbour more than 5.12% LOY-cells, in any of the studied subject. Interestingly, the tools applied for cell clustering identified two separate Treg clusters (Supplementary Figure 2). When further investigated, *CTLA4* expression was found as a major differentiating factor between the Treg clusters. The *CTLA4* gene is expressed on all mature Tregs and is a known regulator of Treg homeostasis^25,26^. Overall, the 8,752 Tregs consisted of 6,975 CTLA4+ and 1,777 CTLA4-cells, and a significant difference in LOY level was found between the two Treg clusters (Wilcoxon signed-rank test: p = 0.00743). The higher frequency of LOY in CTLA4+ cells could suggest that Y loss is influencing the differentiation process of Tregs.

**Figure 2:**
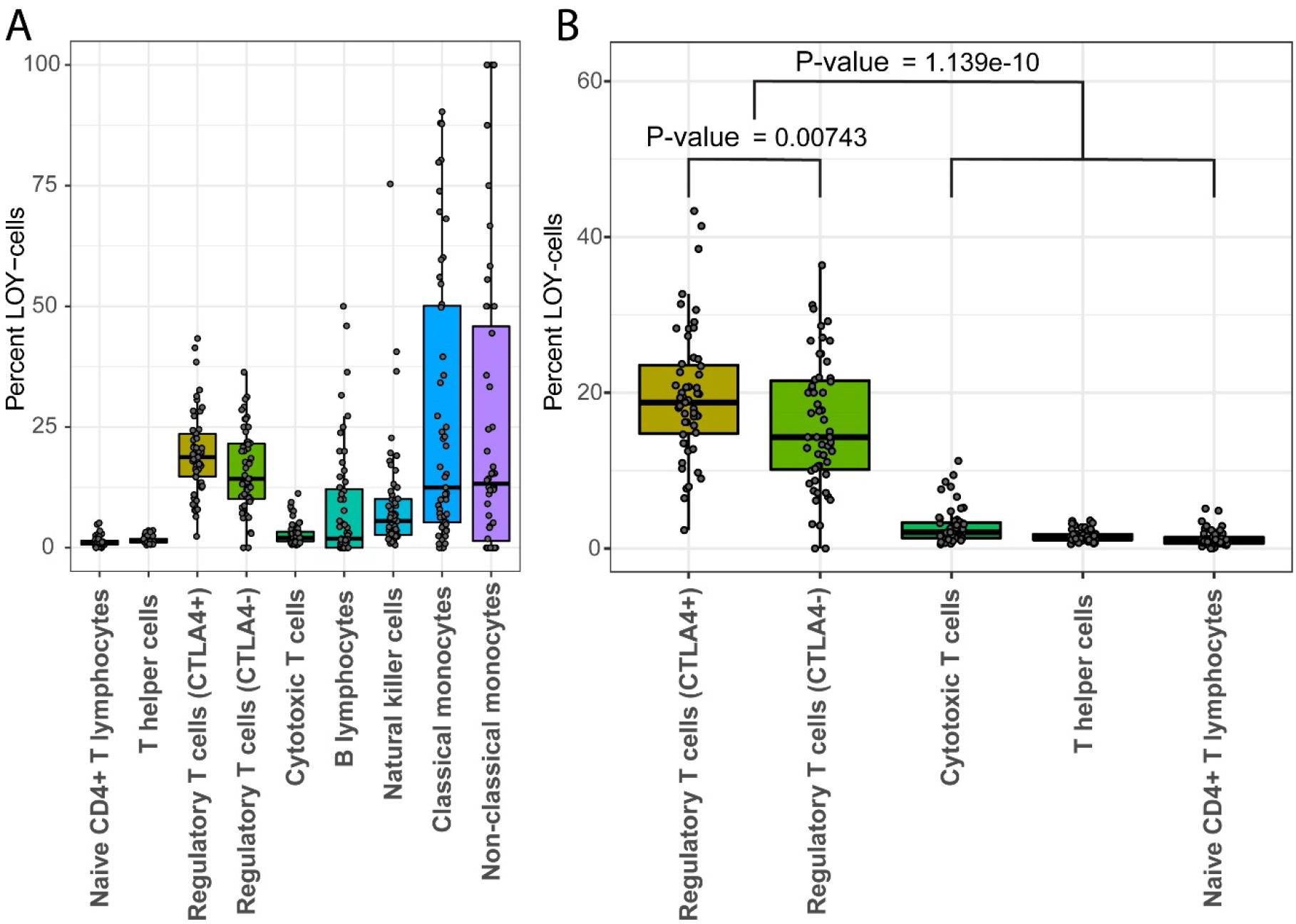
**A)** Boxplot showing the percentage of LOY in the studied leukocytes of the validation dataset. Each dot represents the value of one sample. Median values are marked with black lines, where Tregs have the highest median of any studied cell type. **B**) Subset of *A*; specifically the T lymphocytes.

### LOY as a determinant of CD4+ T lymphocyte cell fate

To replicate the finding from the public datasets that LOY could influence the differentiation of T lymphocytes towards a Treg phenotype, we used a similar approach in the validation dataset. First, linear regression was used to show that the level of LOY in Tregs was positively associated with the fraction of Tregs (Coeff = 0.0394, p = 0.00743). Next, a quasibinomial model was used to establish this result while adjusting for confounders (LR = 5.63, Df = 1 p = 0.0176167). Significant confounders in the model was cell type, whether the sample was CD4+ enriched and interactions between LOY and cell type, as well as cell type and CD4+ enrichment (Supplementary Table 3). Overall, these results independently support the observation from the public datasets that LOY influence Treg abundance. Given this observation, we sought to investigate the hypothesis that Y loss could be affecting the development of CD4+ T lymphocytes by pushing them towards a Treg phenotype. Developmental trajectories for CD4+ T lymphocytes were estimated with pseudotime as a measurement for cell differentiation (Figure 3). The most differentiated CD4+ cells comprised of CTLA4+ Tregs and adjacent T-helper cells, creating a trajectory suggesting a differentiation of naive T lymphocytes with Y loss into CTLA4+ Tregs via the CTLA4-subtype.

**Figure 3:**
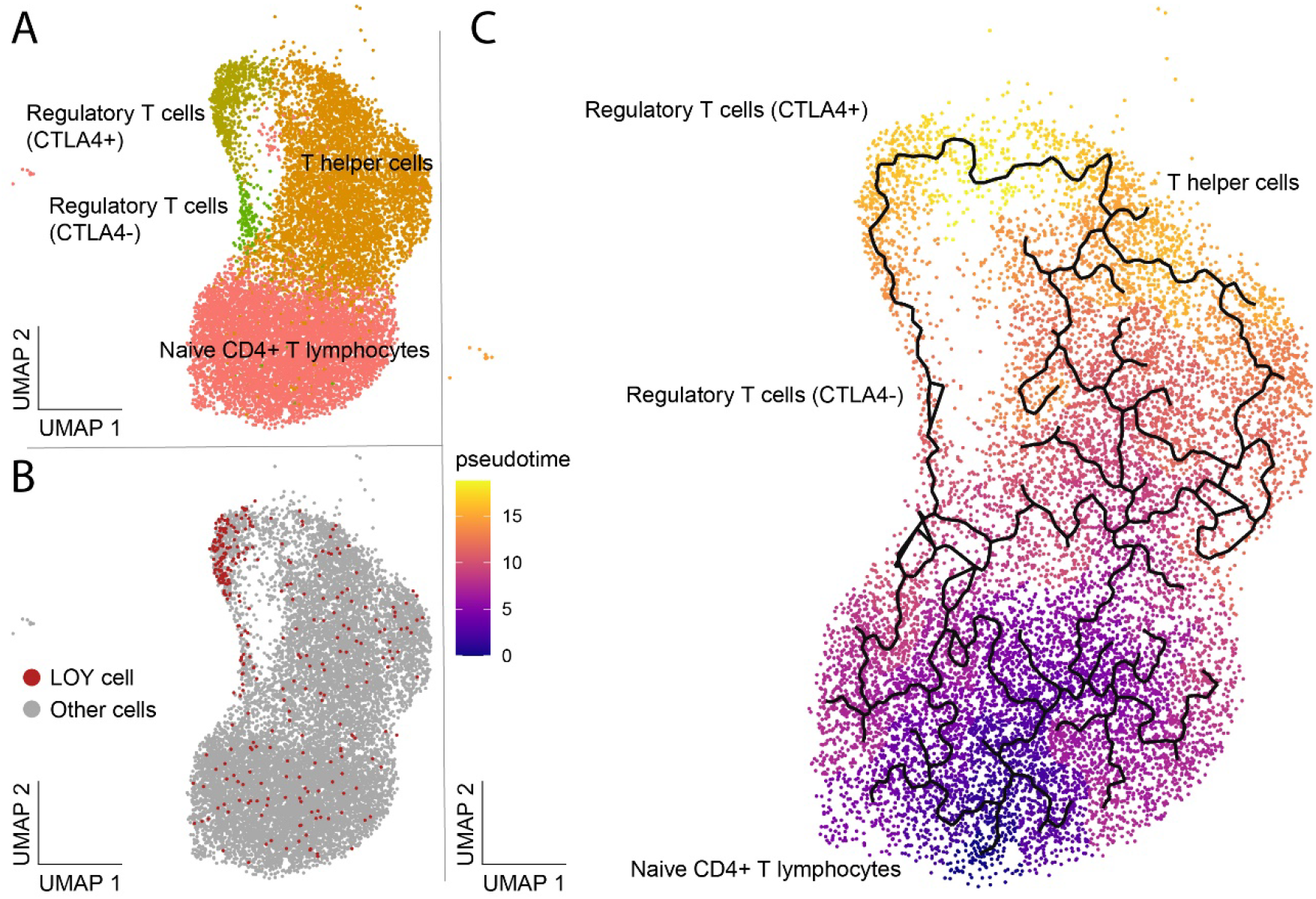
Illustration of the distribution of LOY in different types of CD4+ T lymphocytes, as well as their developmental trajectories. The full UMAP can be seen in *Supplementary Figure 2B*. **A**) The distribution of the identified cell types. **B**) The LOY status of the cells in *A*, with LOY-cells being marked red. **C**) Developmental trajectory of CD4+ T lymphocytes. Colour denotes pseudotime, with more developed cells being brighter. The lines indicate the suggested trajectories from naive to more differentiated CD4+ cells.

### LOY associated transcriptional effects (LATE) in Tregs

Differential expression analysis was used to study changes in autosomal gene expression, referred to as LOY associated transcriptional effects (LATE), in Tregs with Y loss. For the CTLA4-subset of Tregs, only 23 LATE genes were identified (Supplementary Table 4). In contrast, analysis of the CTLA4+ subset of Tregs identified 465 LATE genes, most of which were autosomal genes (Supplementary Table 5). The list of differentially expressed genes contains many that are involved in the normal functions of immune cells, including *S100A11* (logFC = -0.25, adjusted p-value = 8.9e-20), *ANXA1* (logFC = -0.18, adjusted p-value = 4.6e-14), *TIGIT* (logFC = 0.11, adjusted p-value = 0.00011) and *FOXP3* (logFC = 0.10, adjusted p-value = 0.00015). Gene Set Enrichment Analysis (GSEA) found no gene sets for the CTLA4-Treg subset, while the CTLA4+ subset yielded 41 significantly (p < 0.001, Supplementary Table 6) differentiated gene sets. The 20 most significant gene sets from the CTLA4+ analysis were also grouped further into functional categories (Figure 4, Supplementary Figure 5). All significantly upregulated gene sets shared a core of upregulated ribosomal proteins (RPs). In contrast, the downregulated gene sets mainly consisted of genes involved in cell migration and locomotion.

**Figure 4:**
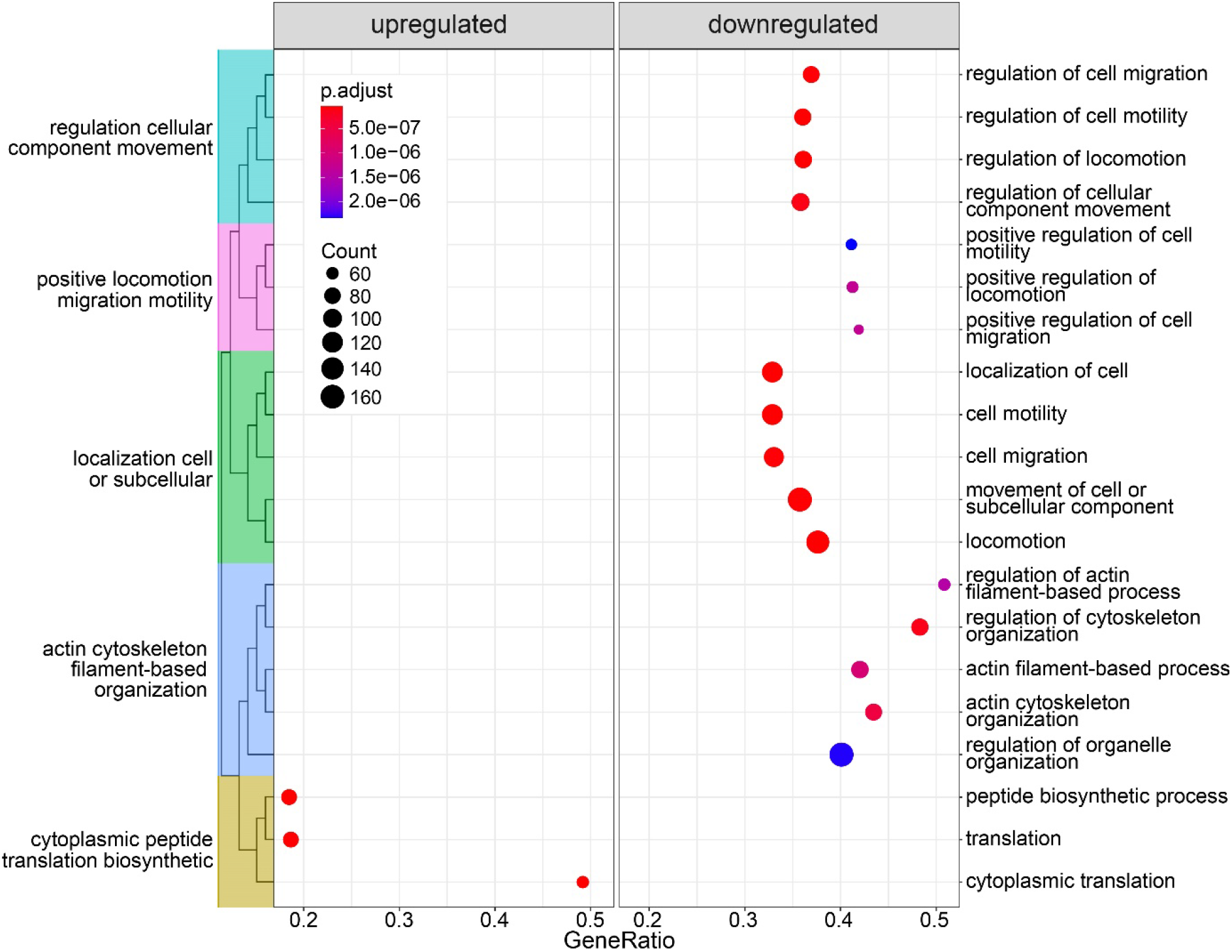
The top 20 categories from the gene set enrichment analysis. Categories within the left and right frames were upregulated and downregulated, respectively. The coloured clusters to the left indicate the same branch of the gene ontology (GO) tree. Dot size and colour indicates gene count and P-value, respectively.

## DISCUSSION

Previous studies have established that LOY is associated with the distribution of different types of blood cells^10-12,14^. Furthermore, it has been shown that the type of leukocyte affected with LOY might be relevant for disease risks, with LOY in specifically CD4+ T lymphocytes being associated with increased risk for prostate cancer^12^. However, LOY occurs at considerably lower frequency in CD4+ T lymphocytes than other leukocytes^3,12,13^. Thus, we investigated here if Y loss disproportionally affects certain subsets of T lymphocytes. In both the public and validation datasets, we found that Tregs had significantly more LOY than any other studied CD4+ T lymphocyte subset. Additionally, higher levels of LOY in Tregs was also positively associated with a higher frequency of Tregs compared with other CD4+ cells. This suggests that Y loss might impact the distribution of T lymphocytes, by pushing naive T lymphocytes towards a Treg phenotype. Alternatively, CD4+ T lymphocytes affected by LOY could undergo apoptosis, with the enrichment for LOY Tregs being due to a lower susceptibility for this apoptosis. However, analysis of developmental trajectories in the validation dataset support the first hypothesis; since it predicts trajectories culminating in the Treg cluster, which has the highest frequency of LOY-cells. Overall, our data suggest that Y loss influence the differentiation of CD4+ T lymphocytes, resulting in an enrichment of Tregs with LOY.

LATE analysis in CTLA4+ Tregs identified 465 dysregulated genes, and the GSEA identified 41 gene sets describing two major effects associated with Y loss. First, the increased expression of RPs identified in LOY Tregs indicates that these cells are upregulating production of ribosomes. Interestingly, losing the Y chromosome involves losing *RPS4Y1*, coding for a ribosomal subunit, and located in the MSY. Its X chromosome homolog, *RPS4X*, escapes X-inactivation, suggesting that the expression of two copies is necessary to maintain dosage^27^. Deletion of ribosomal proteins can activate the mTOR pathway and disrupts ribosomal assembly, resulting in RP upregulation^28^ and transcriptional dysregulation^29^. Thus, the loss of *RPS4Y1* could explain the observed upregulation of RPs, and might contribute to 465 LATE genes found in LOY Tregs. The second major effect identified by the GSEA was several gene sets linked with cell motility. This could be attributed to *CD99*, a gene located in the pseudoautosomal region that will have one copy lost as an effect of Y loss^30^. Studies have previously reported downregulation of *CD99* in LOY-cells, as well as a decreased cell surface abundance of CD99, as an effect of Y loss^12,13^. While a decreased motility of leukocytes with LOY could impair their normal immune functions, it might also influence the varying distribution of LOY-cells in different tissues observed in the public dataset. Here, normal tissue neighbouring tumours was depleted of Tregs and CD8+ T lymphocytes with Y loss, compared with high levels of LOY found in blood and tumour tissue. Tumours are known to recruit suppressive immune cells^19,20^, and it is therefore possible that Tregs with Y loss might be concentrated in the tumour microenvironment due to decreased mobility. In addition to major effects identified by GSEA, certain genes dysregulated in Tregs with LOY should be highlighted. First, both *S100A11* and *ANXA1* genes are downregulated as an effect of Y loss. They encode proteins that forms a complex capable of regulating the EGFR pathway^31-33^, with S100A11 also being a part of the TGF-beta signalling pathway^34^. In specifically T lymphocytes, ANXA1 is conversely an important modulator of proinflammatory functions^35^, with evidence of ANXA1 decreasing the risk of atherosclerosis in humans^36^. *ANXA1* knockout in mouse models lead to chronic inflammation, including lung fibrosis, sepsis, rheumatoid arthritis and atherosclerotic lesion formation^37^. Thus, downregulation of *ANXA1* in Tregs with Y loss could severely limit their normal functions, and in extension inhibit an inflammatory response.

Another interesting gene that is upregulated in Tregs with LOY is *TIGIT*, an immunosuppressive receptor found on tumour-infiltrating NK cells, CD8+, CD4+ and regulatory T cells, with highest abundance in the latter^38^. The TIGIT receptor inhibits immune function by binding with higher affinity to CD155 and CD112 than their usual receptor CD226^38,39^. While the CD226 binding would enhance T lymphocyte and NK cell activation, TIGIT binding instead actively supress immune functions of these cells^40-42^. However, in Tregs, TIGIT is a marker for stability, promoting their immunosuppressive functions further^43^. Additionally, an increased TIGIT to CD226 ratio in the tumour microenvironment has been associated with a higher frequency of activated Tregs, as well as an unfavourable prognosis^44^.

Finally, *FOXP3* was upregulated in LOY Tregs, a major determinant for Treg development and their immunosuppressive activity^45^. Loss of function mutations in *FOXP3* has previously been associated with hyperactive T lymphocytes, as well as fatal immunodysregulation^46^. FOXP3 exerts genome wide regulation of gene expression^47^, including promotion of *CTLA4*^48^, which might be related to the increased level of LOY in the CTLA4+ Treg subset observed here. Since FOXP3 drives Treg development^45^, its upregulation in LOY-cells provides a possible mechanism by which Y loss could influence CD4+ T lymphocyte development.

## CONCLUSION

Here we suggest a possible mechanism to help explain why men with hematopoietic Y loss may have an increased risk of tumour development in other organs. Taken together, our data indicate that LOY could drive the development of CD4+ T lymphocytes towards a regulatory phenotype, leading to enrichment of Tregs with Y loss. Differential expression analysis further highlight genes involved in the immunosuppressive functions of these regulatory cells, potentially linked with the increased vulnerability for cancer previously observed in men affected with LOY.

## Data Availability

All data produced in the present study are available upon reasonable request to the authors

## ACKNOWLEDGEMENTS

We would like to thank the participants. The study nurse Malin Edén at the memory and geriatric clinic, academic hospital for help with the sample collection. The authors Zheng et al., Guo et al., and Zhang et al. for making their scRNAseq datasets publically available. This result is part of a project L.A.F. has received funding from the European Research Council (ERC) under the European Union’s Horizon 2020 research and innovation programme (Grant agreements No. 679744 and 101001789), Additional funding supporting the research to L.A.F.: Swedish Research Council (2017-03762 and 2022-03452), The Swedish Cancer Society (20-1004), Kjell och Märta Beijers Stiftelse, and Konung Gustav V och Drottning Victorias Stiftelse. This study was supported by grants from Swedish Cancer Society, Swedish Research Council (grant number 2020-02010), Swedish Heart-Lung Foundation (grant number 20210051), Hjärnfonden and the Foundation for Polish Science under the International Research Agendas Programme (grant number MAB/2018 /6; co-financed by the European Union under the European Regional Development Fund) to J.P.D.. J.H. received funding from Marcus Borgströms stiftelse. Sequencing was performed by the SNP&SEQ Technology Platform in Uppsala. The facility is part of the National Genomics Infrastructure (NGI) Sweden and Science for Life Laboratory. The SNP&SEQ Platform is also supported by the Swedish Research Council and the Knut and Alice Wallenberg Foundation. The computations and data handling was enabled by resources in project sens2017-134 provided by the National Academic Infrastructure for Supercomputing in Sweden (NAISS) at UPPMAX, funded by the Swedish Research Council through grant agreement no. 2022-06725.

## AUTHOR CONTRIBUTIONS

J.M., J.H., H.D., P.O., M.D., E.R-B., J.P.D. and L.A.F designed the study. J.H., J.P.D. and L.A.F obtained the funding. H.D., B.B-O., P.O., M.D. and E.R-B. performed the experiments. J.M., J.H., J.B., A.L., A.Z. and L.A.F analysed the data and interpreted the results. H.D., B.B-O. and J.P.D. contributed to sample collection. J.M., J.H. H.D., J.B. and L.A.F wrote the first version of the paper. All authors contributed to editing the final version of the paper.

## METHODS

### Collection and sequencing

The patient cohort selected for enrichment of CD4+ leukocytes included 30 male participants from the Epidemiology for health study (EpiHealth) and 2 males from the Uppsala-Umeå Comprehensive Cancer Consortium (UCAN). 32 ml of blood was collected into four BD Vacutainer® CPT™ Mononuclear Cell Preparation Tubes (BD), and PBMCs were isolated following the manufacturer’s instructions. The PBMCs were then washed with PBS and cell number and viability were estimated with EVE™ Automated Cell Counter (NanoEnTek) using trypan blue. CD4+ T lymphocytes were enriched from the PBMCs using CD4+ T Cell Isolation Kit human (Miltenyi Biotec) according to the manufacturer’s instructions. Enriched CD4+ T-cells were then diluted to a concentration of 10^6^ cells/ml in PBS with 0.04% BSA with a cell viability of >90%. scRNAseq libraries of CD4+ T-cells were generated using Chromium Next GEM Single Cell 3’ Reagent kit v3.1 (10x Genomics) according to the manufacturer’s instructions. The single-cell libraries were then sequenced using the NovaSeq 6000 and v1.5 sequencing chemistry (Illumina Inc.). The single-cell library preparation and sequencing were performed at the Science for Life technology platform SNP&SEQ, Uppsala University, Sweden. Additionally, 23 scRNAseq datasets derived from PBMCs from the Uppsala Alzheimer’s Disease cohort (UAD) were used as validation. Sample preparation was the same as above except for the CD4+ T Cell enrichment. Overview of participants and their clinical characteristics can be seen in Supplementary Table 7. Informed consent was obtained from all the participants, or from next of kin. The research was approved by the local research ethics committee in Uppsala, Sweden Dnr. 2015/458 (EpiHealth cohort), amendment Dnr. 2015/458/2 (UCAN cohort), Dnr. 2015/092 (UAD cohort).

### Pre-processing, mapping and LOY cell identification

Each sequenced sample was mapped using the Cell Ranger pipeline (v. 6.0 10X Genomics) and standard settings. Following this, the velocyto software was used, counting reads from expressed transcripts and also counting intronic reads as well as reads in the untranslated regions. Generated count matrices from both software were read into an R environment for further study. To identify cells with LOY, all generated count matrices were used and cells showing no reads mapping to the male specific Y were considered to be LOY cells. In contrast to previous methods, this included any MSY reads identified by velocyto as well as cell ranger. This was benchmarked using an established method recommending that each cluster has a score of at least 250 expression from MSY to call LOY-status^24^. The new approach scored higher than 250 in all clusters (Supplementary Figure 3), which would not be the case with previous methods (Supplementary Figure 4).

### Data harmonization and quality control

Unless specified otherwise, the following analysis steps were performed in R version 4.0.4, using Seurat version 4.0.1, on the UPPMAX Bianca computational cluster. Two criteria were used to exclude low quality cells, the number of expressed features and the percent mitochondrial reads. Cells were required to have more than 500, but less than 3000 expressed features, as well as less than 13% of all reads being mitochondrial. These thresholds were chosen based on the variable distribution (Supplementary Figure 6). The count matrix was thereafter normalized using SCTransform (version 0.3.2), regressing out effects based on percent mitochondrial reads. The samples were thereafter integrated, based on 3000 features and one sample (SF-2212-EPH001) as reference. The integration functions were chosen to account for the SCTransform. In addition to SCTransform, log-based normalization was also done using Seurats NormalizeData function with default settings. The log-based normalization is preferred when investigating differentially expressed genes, as advised by the team behind Seurat. After calculating principal components, batch effects introduced by sampling and sorting were removed using Harmony (version 1.0). The number of harmonized principal components to use for clustering was thereafter chosen based on an Elbowplot (Supplementary Figure 7). Clustering was performed on the 22 first principle components with 0.9 resolution.

### Cell type identification

The classification of cell type identity was guided by two tools, CIPR (version 0.1.0) and singleCellNet (version 0.1.0). CIPR was run on marker genes, calculated using the FindAllMarkers function on log-normalized data. The Presorted PBMC single-cell RNAseq dataset, hsrnaseq, was used as reference. The resulting CIPR classification can be seen in Supplementary Figure 8A. SingleCellNet was trained using data from Zheng et al.^49^, available through the 10X Genomics Datasets database. The training dataset was filtered (200 < number of features < 1500 & percent mitochondrial < 5%) and normalized according to the main dataset. It was then trained (nTopGenes = 10, nRand = 70, nTrees = 1000, nTopGenePairs = 25) and prediction scores tested (nrand = 50). Thereafter, singleCellNet classification was run (nqRand = 50) on the log-normalized main data. The singleCellNet classification can be seen in Supplementary Figure 8B, where the most commonly predicted cell type per cluster was used as identity for the entire cluster. The cell identities suggested by each tool were used to guide cell type classification, which ultimately was decided based on the expression of known marker genes in each cluster. These known marker genes included *CD4, FHIT* and *CCR7* for Naïve CD4+ T lymphocytes, with *FHIT* and *CCR7* negative as Helper T cells. *CD4, FOXP3, IL2RA* and *TIGIT* for Tregs, which were further separated by *CTLA4. CD8* for Cytotoxic T cells, while *NKG7* and *GNLY* indicated NK cells. Classical monocytes were identified by *FCN1* and *CD14*, with non-classical monocytes defined by the addition of *FCGR3A*. B-lymphocytes by *CD19* and *VPREB3*. Additional clusters, exhibiting gene expression profiles not indicating any of the above marker genes, were classified as unidentified.

### Differential expression analysis

Genes differentially expressed due to LOY were found with the Limma-trend algorithm. This was done per cell type, and included genes expressed in at least 10% of cells in the studied cell type. Using the Limma R package (version 3.46.0), the model matrix was defined with LOY status and sample origin specified as covariates. The model was thereafter fitted on the Log-normalized expression data with default settings, as well as LOY status set as the coefficient of interest and Benjamini-Hochberg as p-value adjustment method.

### Gene set enrichment analysis

Fold changes for all genes, calculated using the limma package (see differential expression analysis), was collected. Genes on the male specific region of the Y chromosome were removed. After this, clusterProfiler (version 4.2.2) was used with fold changes for each gene, tested by the limma package, as a metric to calculate the enrichment of all gene sets present in the “Biological process” category of the gene ontology resource (http://geneontology.org/). Clustering for categories was performed using the pairwise_termsim function from the enrichplot R package (version 1.13.1.992)

### Developmental trajectories

The cells defined as Naïve CD4+ T lymphocytes, Helper T cells and Tregs were selected to estimate developmental trajectories. To avoid an issue with overfitting, the number of cells was randomly decreased by a factor of 10 using the sample function in R. The seed was set to 14 for this step. The SeuratWrapper package (version 0.3.0) was thereafter used in R (version 4.1.3) to transform the sampled SeuratObject into a CellDataSet object used by Monocle3 (version 1.0.0), which was used to predict developmental trajectories. Monocle3 was run on the UMAP previously constructed, using the Louvain clustering method. The cluster designated as Y_73 was thereafter used as the root. Prior to running the developmental trajectory analysis, the random generator seed was set to 1477.

### Public dataset

The three public datasets were collected via their corresponding GEO catalogs, GSE98638, GSE99254 and GSE108989. They were processed similarly to as described above, with the exception of the steps described here. The analysis was performed in R version 4.1.3 with Seurat version 4.1.0. Any step prior to integration was performed independently for each dataset. Firstly, low quality cells were filtered based on the number of expressed features (nFeats) and UMI count (nUMI). nFeats was used instead of percent mitochondrial read as this was not available for the public datasets. The thresholds were chosen based on the corresponding distribution (Supplementary Figure 9) for liver cancer (2300 < nFeats < 4400; 3e5 < nUMI < 1.2e6), lung cancer (1800 < nFeats < 5200; 2e5 < nUMI < 1e6) and colorectal cancer (1800 < nFeats < 5200; 2e5 < nUMI < 1e6). Metadata was thereafter collected for the cells that passed filtering via the identifier assigned to each cell by the original authors. Except for the numbers identifying the patient of origin for each cell, combinations of letters could be used to discern the sampled tissue and sorting. In addition to P, T and N denoting peripheral blood, tumor tissue and normal adjacent tissue, TR, TC and TH identified Tregs, cytotoxic T cells and T helper cells, respectively. Other identifiers were not present in all three datasets; these cells were therefore excluded. To classify LOY-status, the sex of each original sample was identified by comparing the expression of MSY genes. The list of Y located genes was collected using the BioMart package (version 2.40.0) on Ensembl (version 99). While the sex of each patient was clear, some females cells still expressed MSY genes. Considering the female MSY expression background noise, a threshold was created at the 95th quantile of total MSY expression in specifically female cells with any MSY expression. Additionally, most female MSY expression was from a single MSY gene. Thus, any male cells with a total MSY expression less than this threshold, as well as expression from only one or less MSY gene, were classified as LOY (Supplementary Figure 10). After normalization using SCTransform (version 0.3.3). The three datasets were integrated, followed by the calculation of principal components. Harmony was used to remove batch effects attributed to patients and tissue of origin.

The first 15 dimensions from Harmony, chosen based on elbow plot (Supplementary Figure 11), were finally used to cluster the cells to a resolution of 0.6. Due to the poor performance on this dataset by both cell type classification algorithms, the cell type of each cluster was manually designated based on only the expression of known marker genes.

### Statistics

To test if LOY was more common in Tregs than other T lymphocytes, Wilcoxon signed-rank test was used to compare the difference between LOY percentage values from the sample. For the validation dataset this was first done between Tregs and other T lymphocytes to reduce the number of tests performed, thereafter testing the difference between the Treg subsets. When comparing the Treg versus other CD4+ T lymphocytes abundance, non-parametric linear regression was used via the mblm package (version 0.12.1). A non-parametric model was necessary as the variables were not normally distributed. Further models to test the association between LOY-level and other factors such as cell type and tissue was run either as an quasibinomial model with R’s glm function or a negative binomial model run with the MASS package (version 7.3-55). The negative binomial model was used for the LOY to cell type test in the public datasets to account for additional data structures and sorting. Quasi models was also necessary to handle high levels of residual deviance. The produced models were run as a type 3 ANOVA using the car package (version 3.0-10 and version 3.0-12 for the validation and public datasets, respectively), with contrasts defined as options(contrasts = c(“contr.sum”, “contr.poly”)). See *Supplementary Table 1-3* for full models.

