## Supplemental material for "Loss of chromosome Y in regulatory T cells"

### Supplementary Figures

#### Supplementary Figure 1

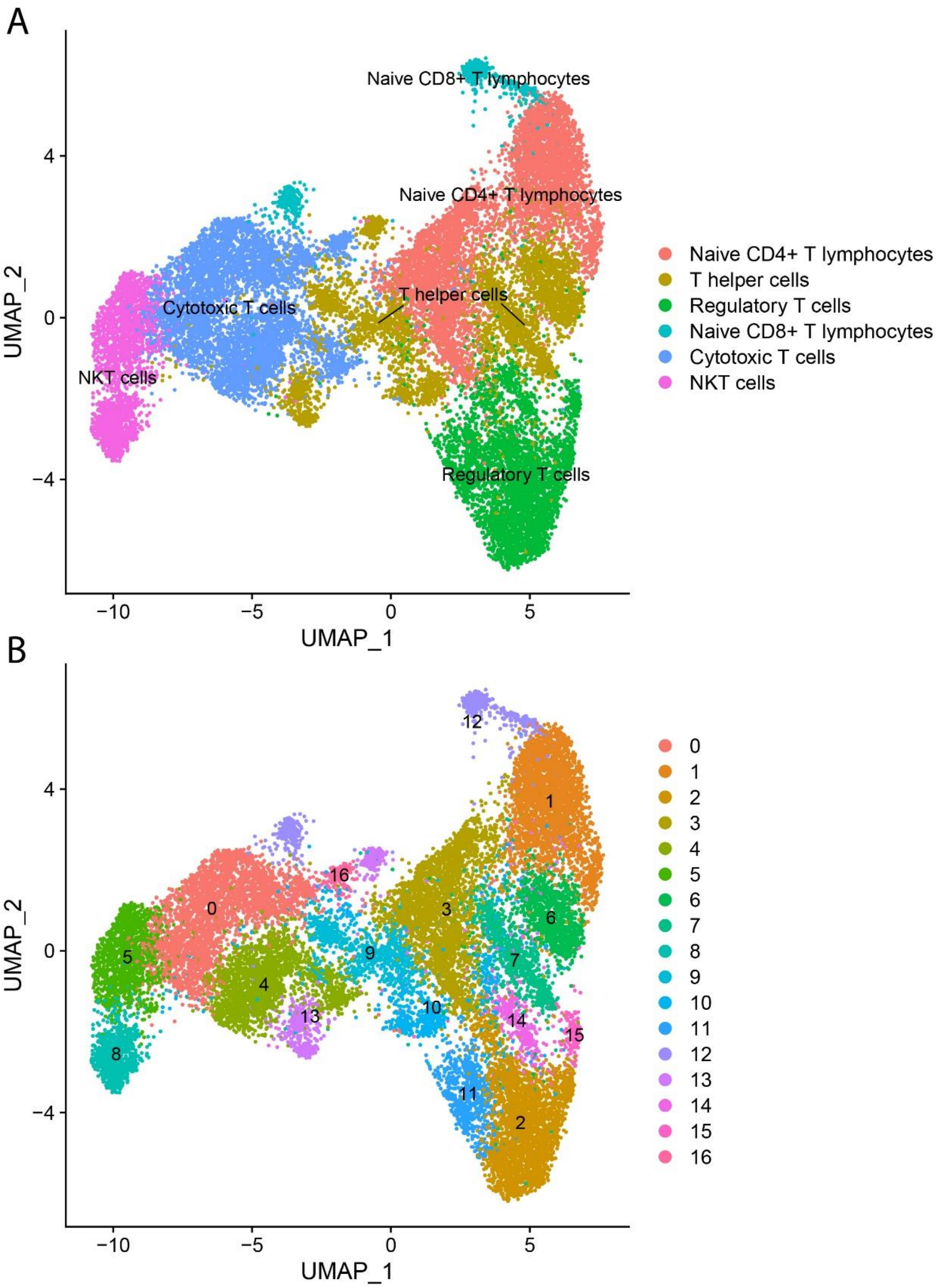


**Supplementary Figure 1**: UMAP from the public scRNAseq datasets, calculated on 15 principle components, showing **A**) the assigned cell types and **B**) clustering of cells with 0.6 resolution.

Supplementary Figure 2

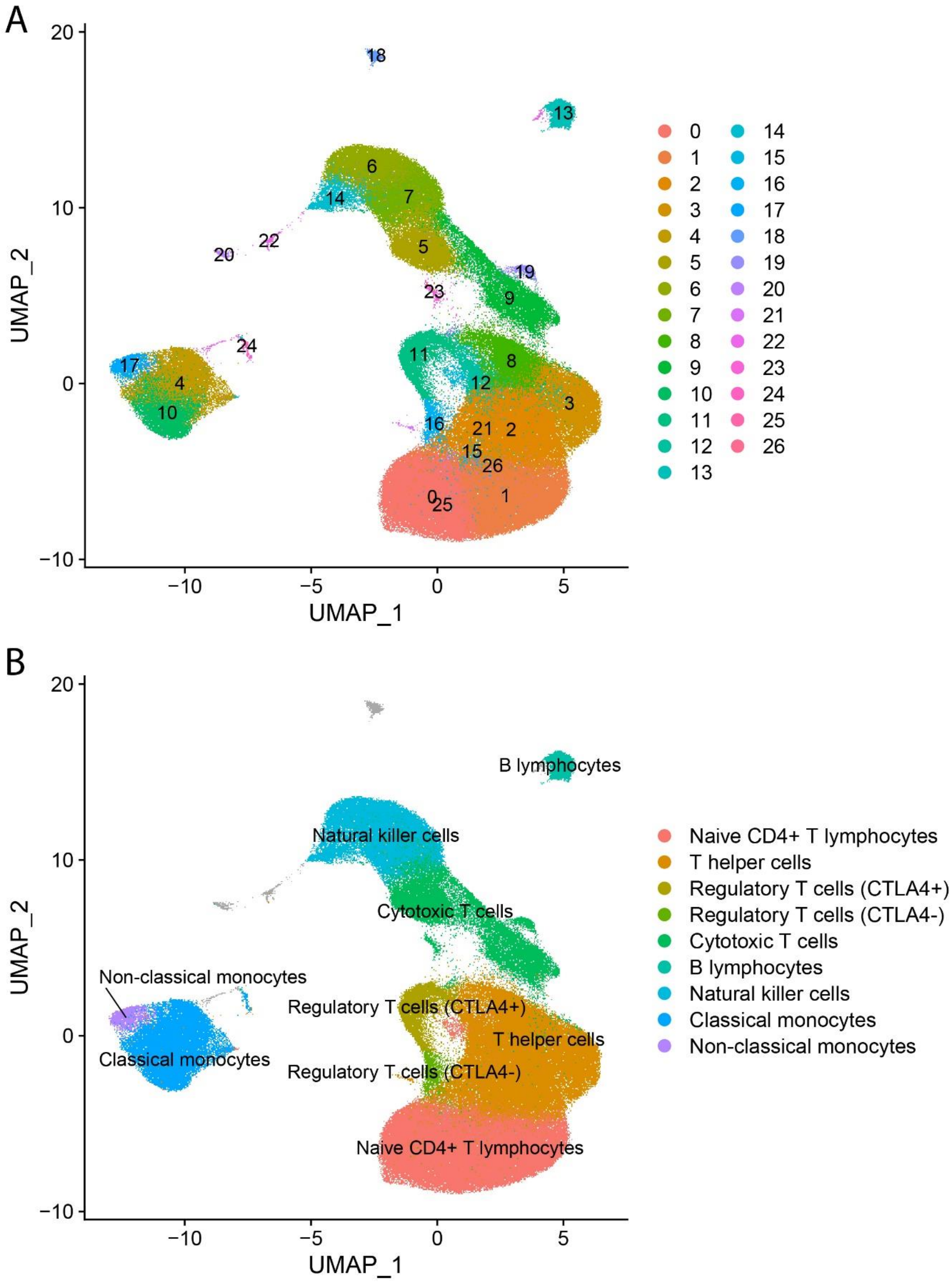


**Supplementary Figure 2**: UMAP from the validation scRNAseq datasets calculated on 22 principle components showing **A**) the clustering of cells with 0.9 resolution and **B**) the assigned cell types. Grey marks cells within clusters not identified as any of the studied cell types.

Supplementary Figure 3

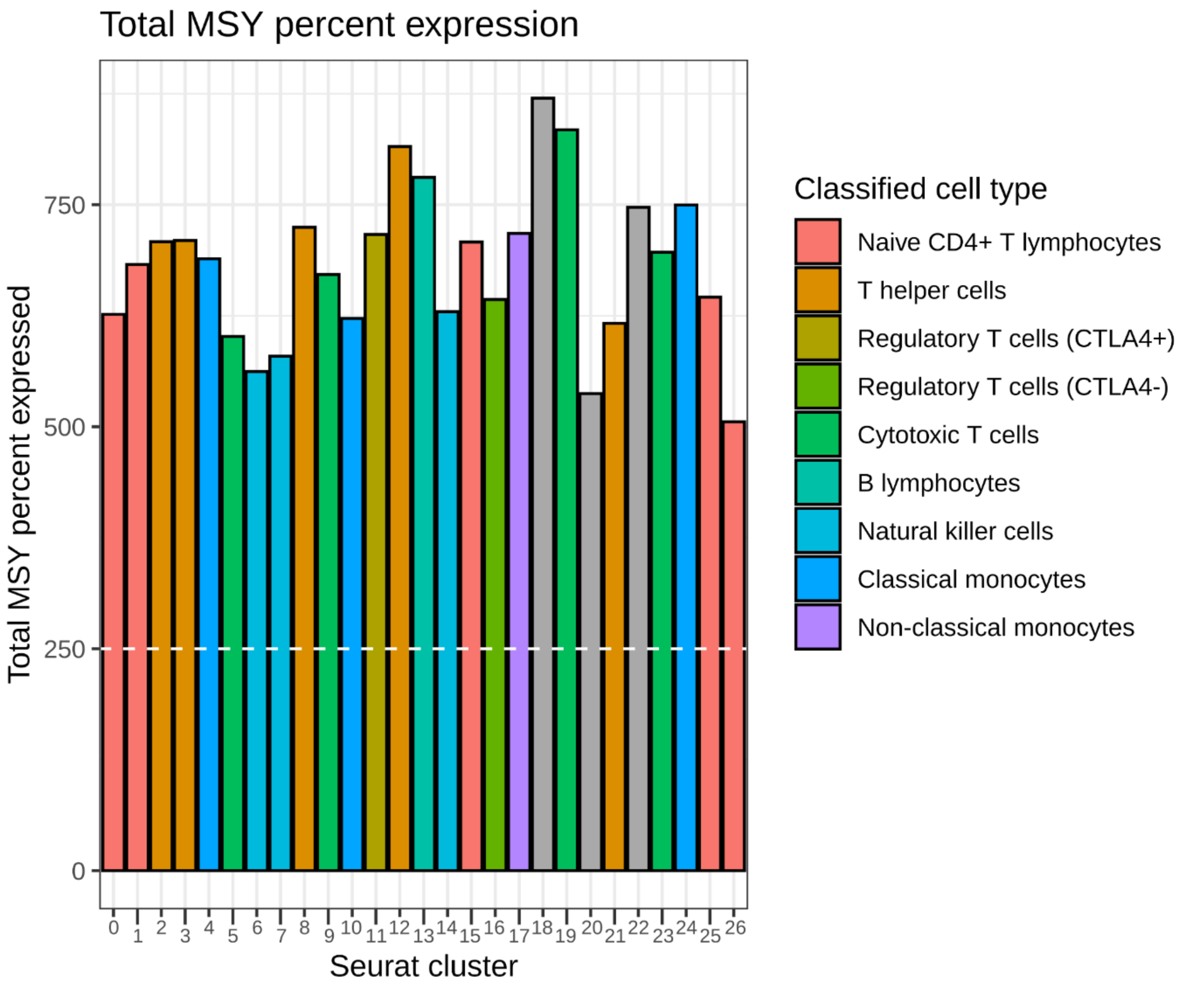


**Supplementary Figure 3**: Barplot showing the total cumulative MSY percent expression in each cluster, a method devised to qualify LOY assessment. The dashed line shows a cutoff at 250, the published QC threshold.

Supplementary Figure 4

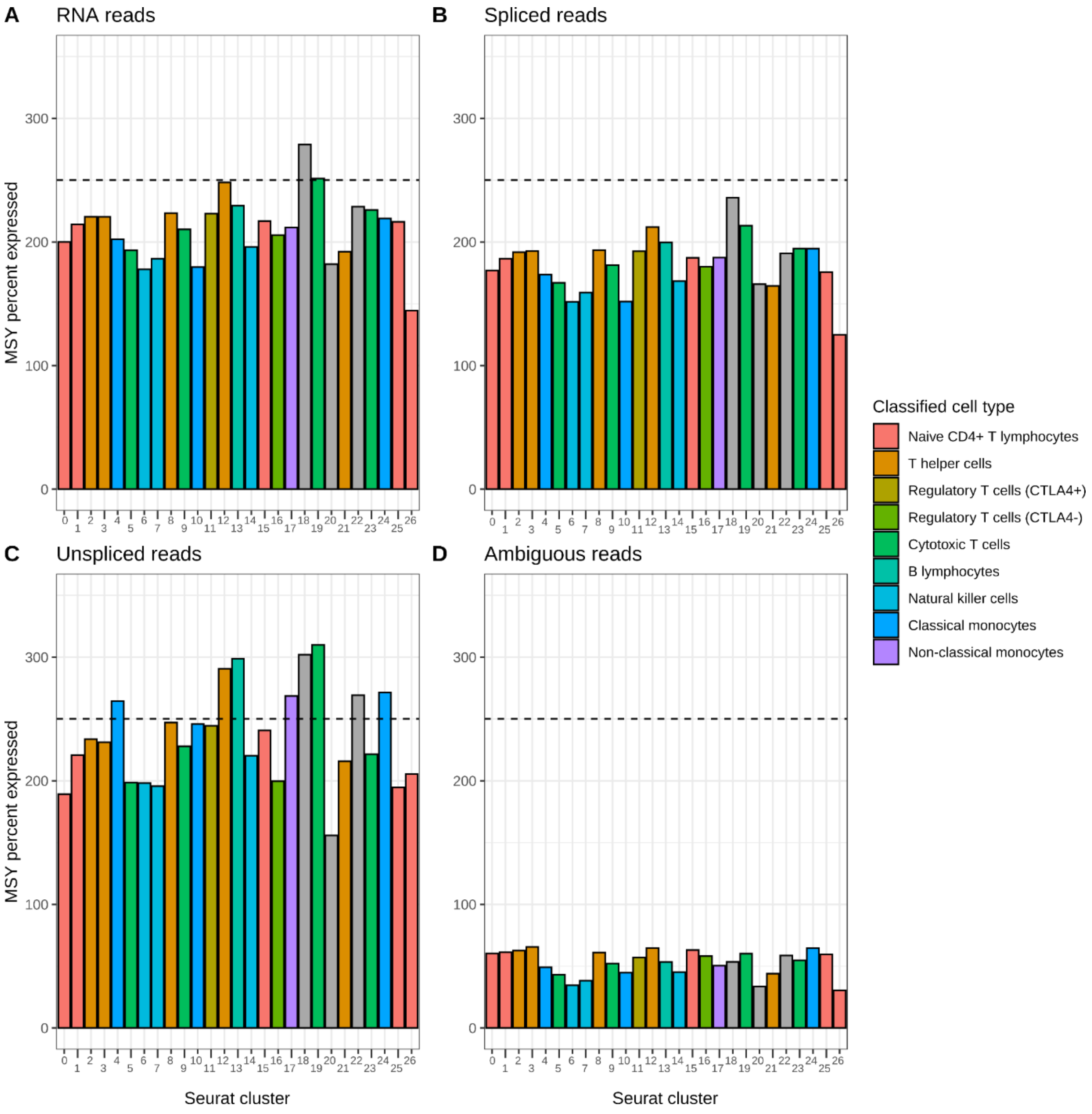


**Supplementary Figure 4:** The Barplot in *Supplementary Figure 3* separated by assay used to estimate LOY. Whereas the previous method of LOY classification only used the RNA reads identified by Cell Ranger (**A**), the new method considers all the different assays (**A**-**D**). The dashed line shows the published threshold required for LOY scoring.

Supplementary Figure 5

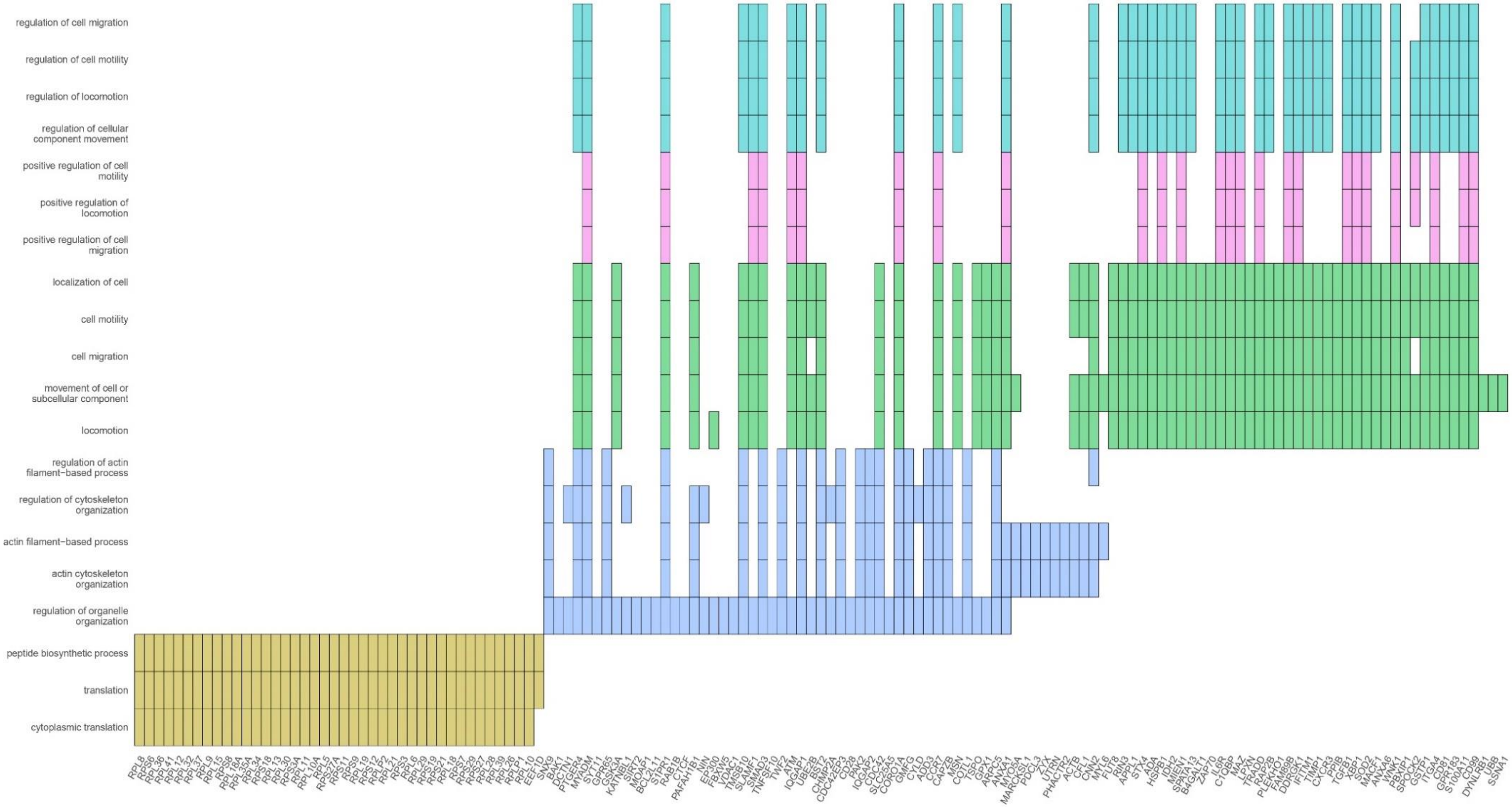


**Supplementary figure 5:** Inclusion of expressed genes in the different significant categories presented in *Figure 4*.

Supplementary Figure 6

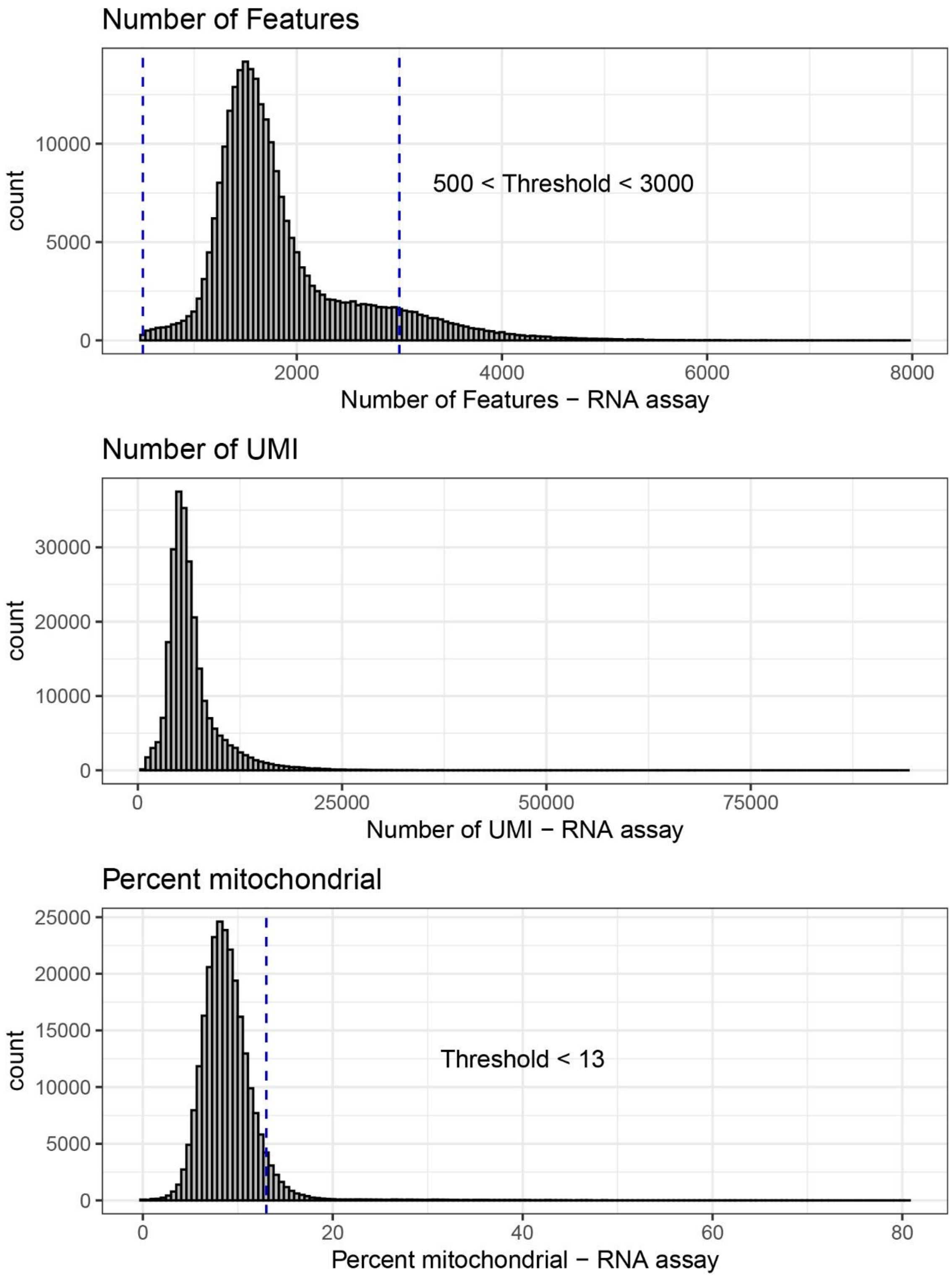


**Supplementary Figure 6**: Histograms presenting the filtering threshold used for the validation dataset. For each QC variable, the dashed line indicate the threshold applied.

Supplementary Figure 7

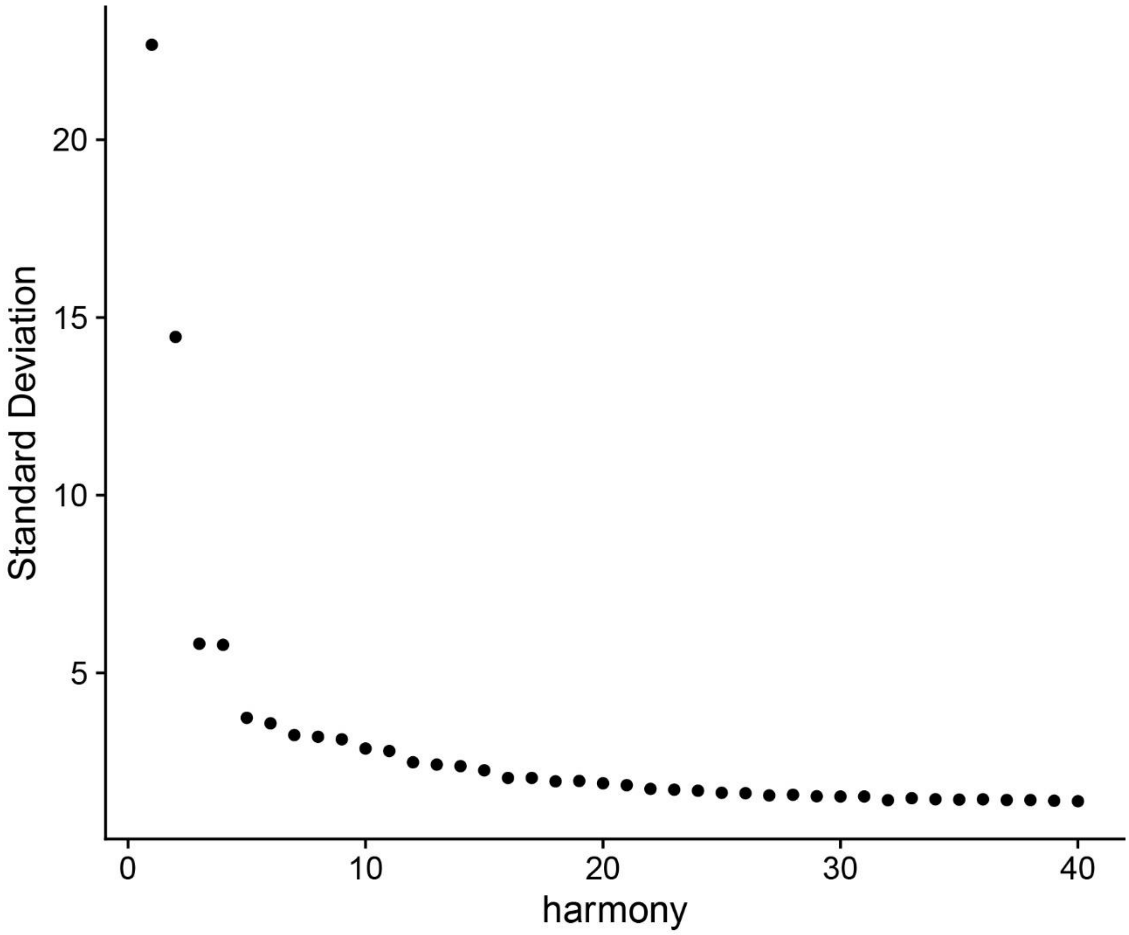


**Supplementary Figure 7**: Elbow plot used to pick the number of principle components used for clustering for the validation dataset. Based on this figure, the first 22 principle components were chosen.

Supplementary Figure 8

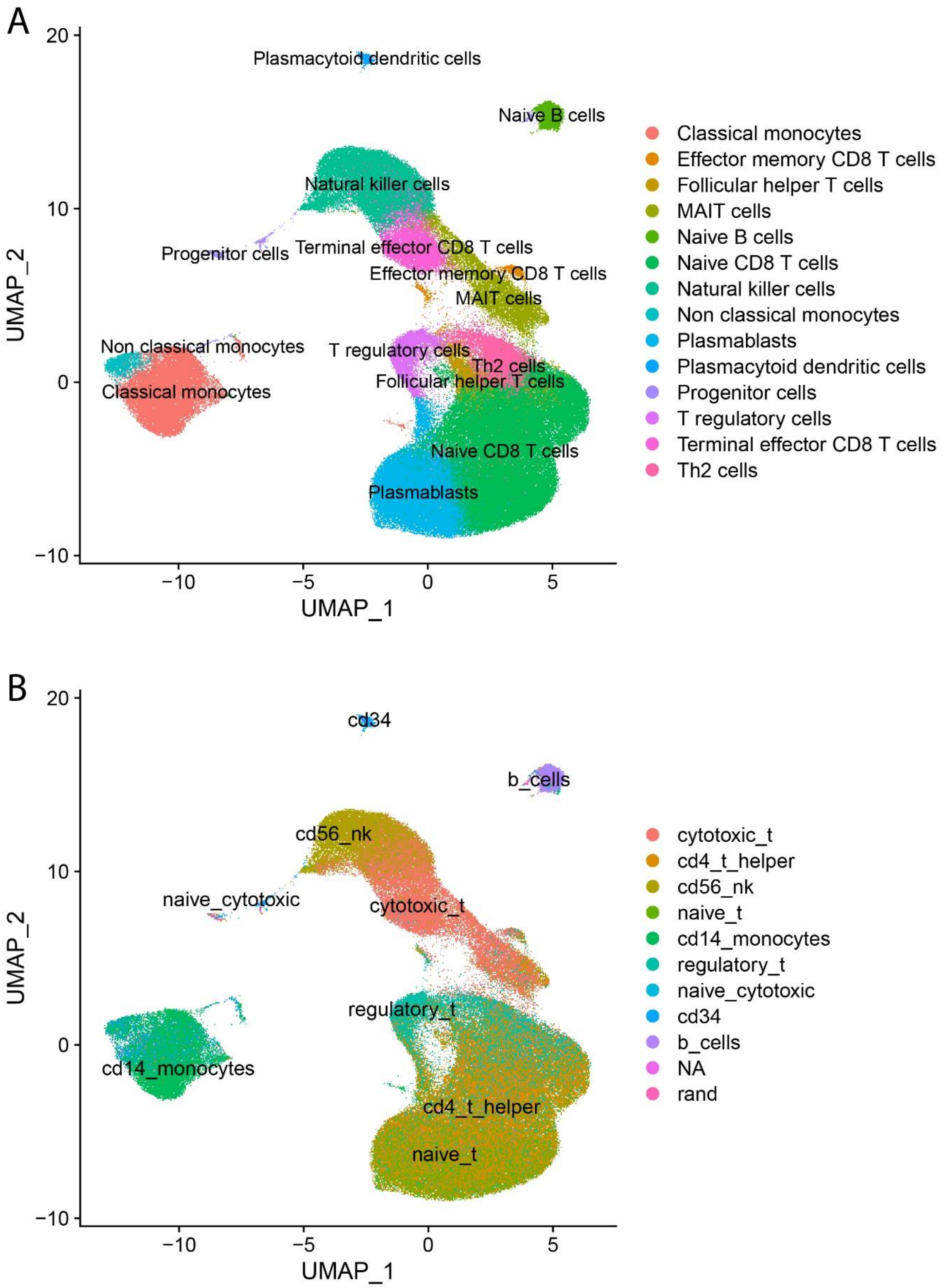


**Supplementary Figure 8**: UMAP showing the cell types predicted by the classification tools **A**) CIPR and **B**) SingleCellNet. This was used to guide the final cell type classification (*Supplementary Figure 2B*).

Supplementary Figure 9

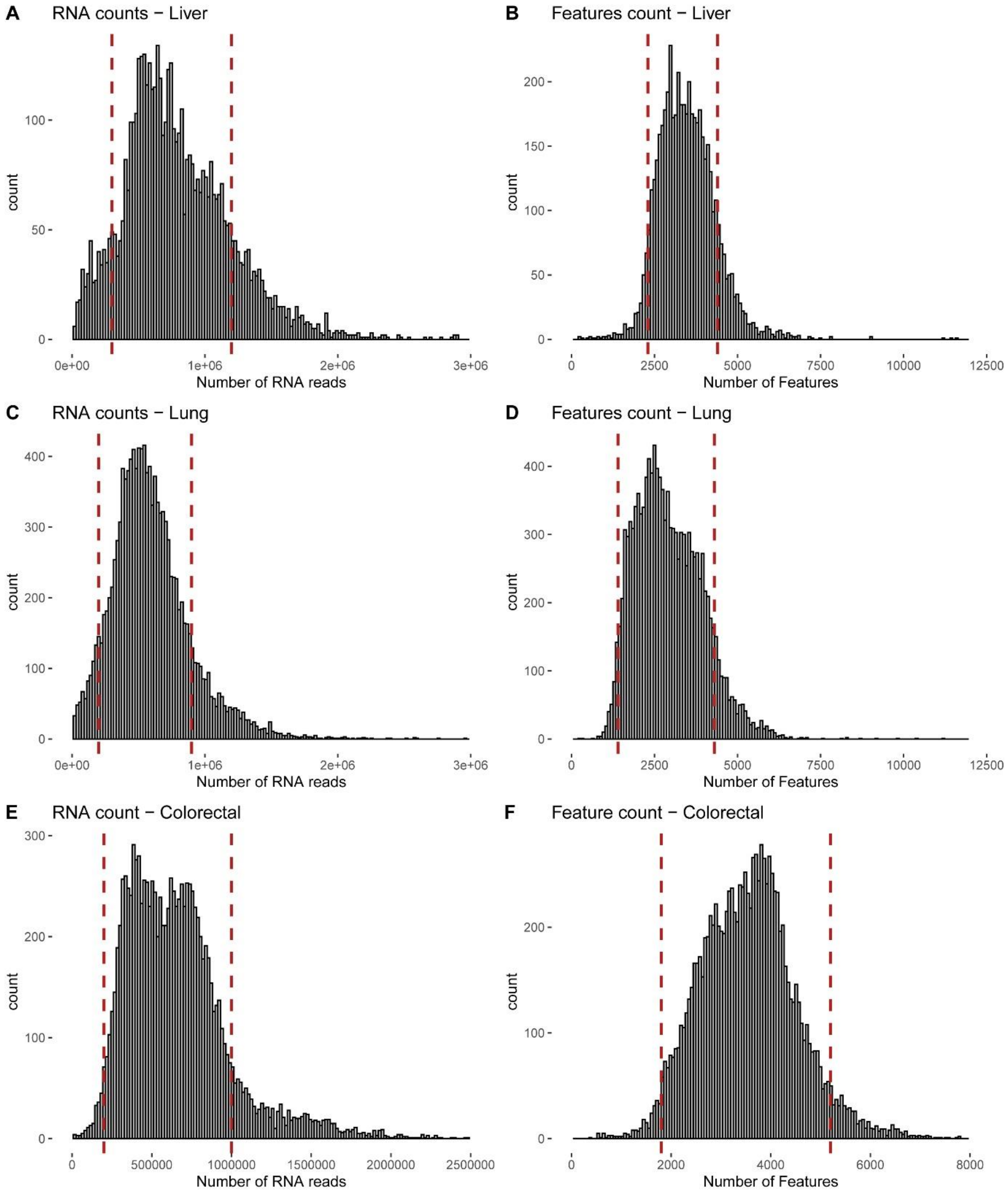


**Supplementary Figure 9**: Histograms presenting the filtering threshold used for the public datasets, similar to *Supplementary Figure 6*. Each dataset (here shown by row) was QC independently based on both RNA count and Feature count (by column).

Supplementary Figure 10

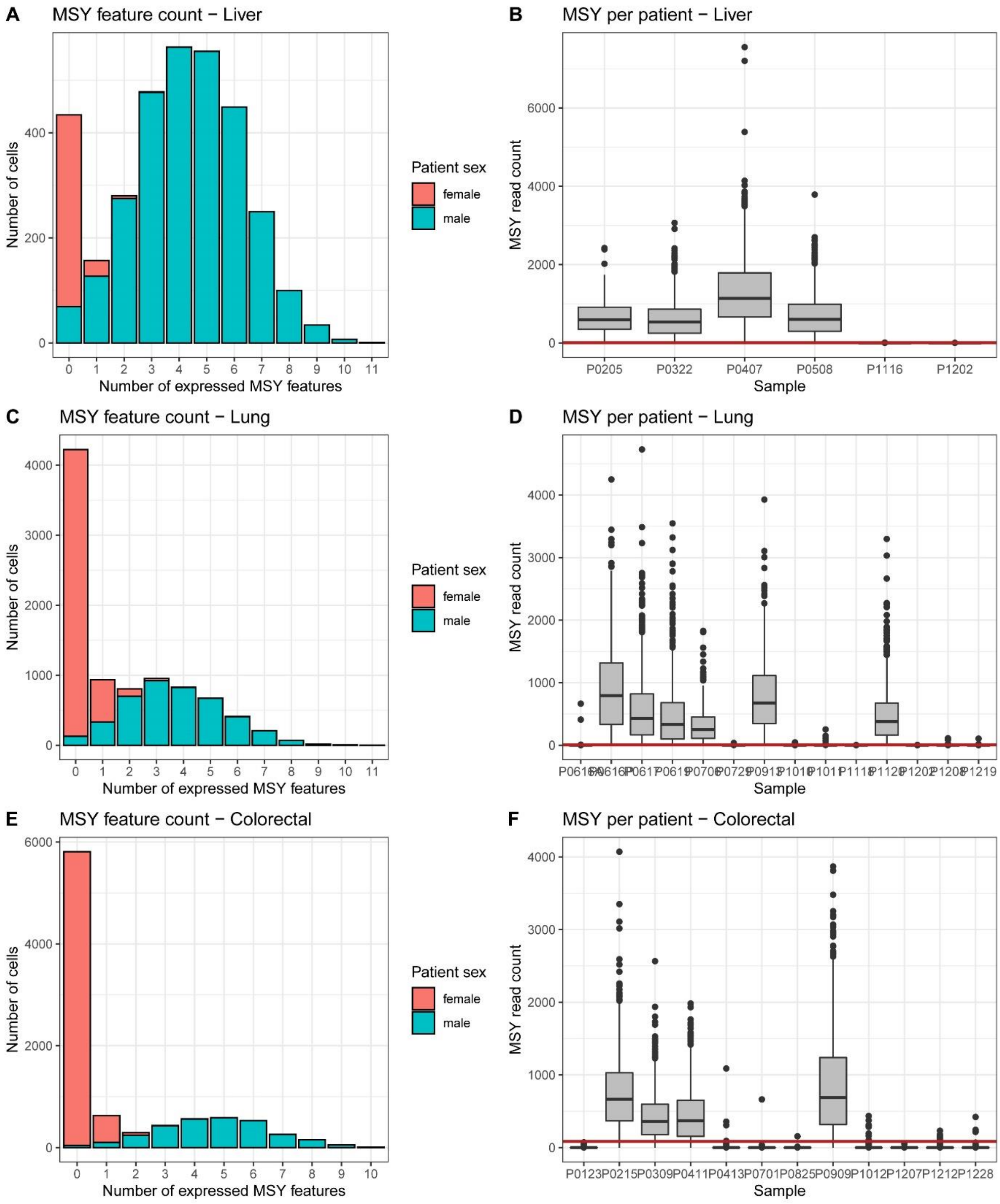


**Supplementary Figure 10**: Plots used to determine the method and threshold of LOY classification in the public dataset. This was done independently for each dataset, here shown by row.

Supplementary Figure 11

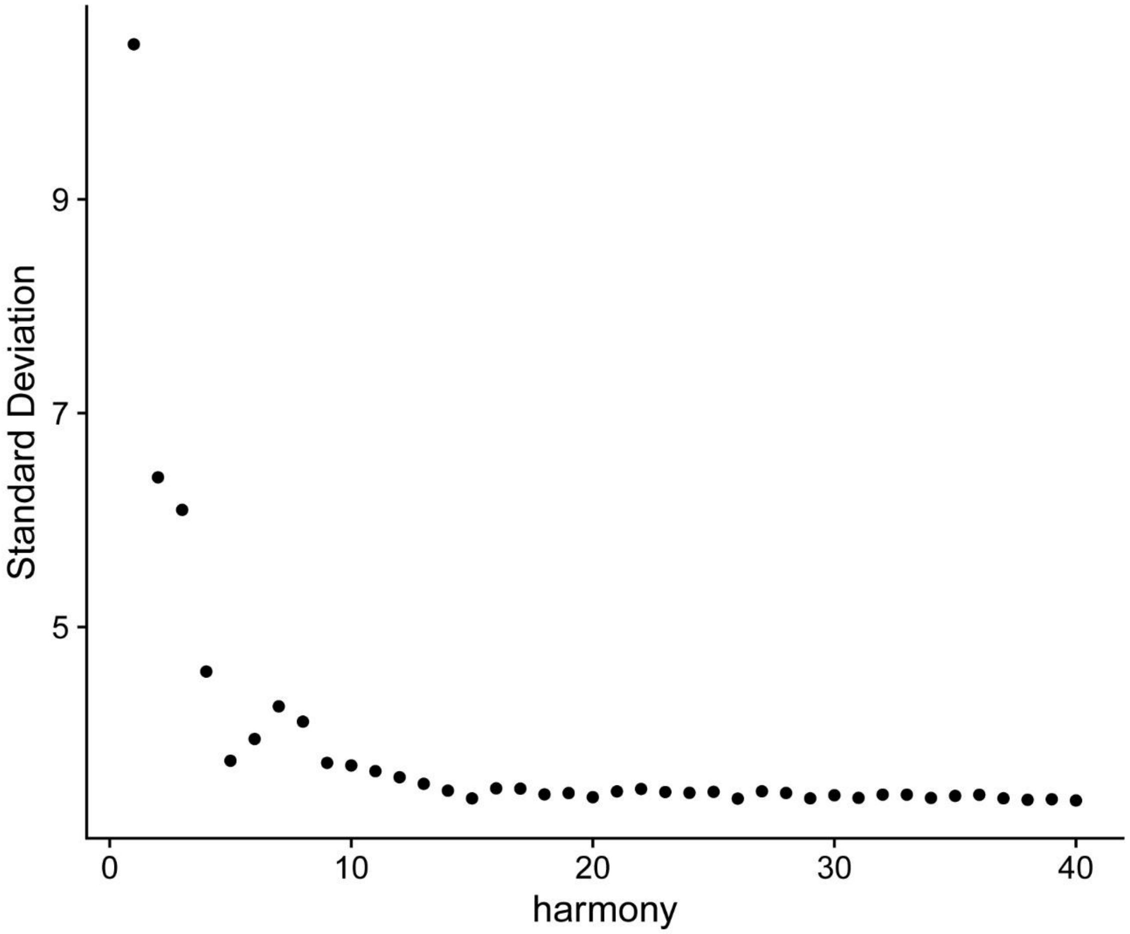


**Supplementary Figure 11**: Elbow plot used to pick the number of principle components used for clustering for the public datasets. Based on this figure, the first 15 principle components were chosen.

### Supplementary Tables

**Supplementary Table 1**: Statistical model to test whether the number of cells in each cell type and sample depends on LOY. Type III ANOVA based on a negative binomial model. Logarithmic link function. Cell count was used as response variable. Deviance of 89.10299 on 58 residuals.

| | LR Chisq | Df | Pr(>Chisq) |
|---|---|---|---|
| scale(percent.LOY) | 6.591 | 1 | 0.01025 * |
| cell.type | 81.317 | 5 | 4.45e-16 *** |
| cancer.type | 2.391 | 2 | 0.30263 |
| scale(percent.LOY):cell.type | 3.394 | 5 | 0.63941 |
| cell.type:cancer.type | 14.213 | 10 | 0.16348 |
| scale(percent.LOY):cancer.type | 8.423 | 2 | 0.01483 * |

**Supplementary Table 2**: Statistical model to test whether proportion of LOY cells is significantly different between tissues. Type III ANOVA based on a quasibinomial model. Logistic link function. Proportion of LOY cells compared with normal cells used as response variable. Deviance of 335.8531 on 199 residuals.

| | LR Chisq | Df | Pr(>Chisq) |
|---|---|---|---|
| cell.type | 64.094 | 5 | 1.727e-12 *** |
| tissue | 8.426 | 2 | 0.014804 * |
| cancer.type | 13.436 | 2 | 0.001209 ** |
| tissue:cancer.type | 16.326 | 4 | 0.002611 ** |
| cell.type:cancer.type | 22.680 | 10 | 0.011990 * |

**Supplementary Table 3**: Statistical model to test whether the proportion of cells in each cell type and sample depends on LOY. Type III ANOVA based on a quasibinomial model. Logistic link function. Proportion of cells in current cell type compared with other cell types used as response variable. Deviance of 38355.77 on 616 residuals. Batch denotes sample type, either CD4+ T lymphocyte enrichment or PBMC.

| | LR Chisq | Df | Pr(>Chisq) |
|---|---|---|---|
| scale(percent.LOY) | 5.63 | 1 | 0.0176167 * |
| cell.type | 810.08 | 11 | < 2.2e-16 *** |
| batch | 14.85 | 1 | 0.0001166 *** |
| scale(percent.LOY):cell.type | 22.73 | 11 | 0.0192874 * |
| scale(percent.LOY):batch | 1.75 | 1 | 0.1864470 |
| cell.type:batch | 617.53 | 11 | < 2.2e-16 *** |

**Supplementary Table 6**: The results from the GSEA. Only gene sets significant after adjusting for multiple testing (Benjamini-Hochberg) was included.

| ID | Description | setSize | enrichmentScore | NES | pvalue | p.adjust |
|---|---|---|---|---|---|---|
| GO:0002181 | cytoplasmic translation | 122 | 0.64020 | 3.02877 | 1.00E-10 | 4.31E-08 |
| GO:0006412 | translation | 402 | 0.31803 | 1.66259 | 1.00E-10 | 4.31E-08 |
| GO:0040011 | locomotion | 396 | -0.50748 | -1.66080 | 1.00E-10 | 4.31E-08 |
| GO:0040012 | regulation of locomotion | 241 | -0.55807 | -1.80073 | 1.00E-10 | 4.31E-08 |
| GO:0043043 | peptide biosynthetic process | 406 | 0.31457 | 1.70744 | 1.00E-10 | 4.31E-08 |
| GO:0048870 | cell motility | 365 | -0.51881 | -1.69465 | 1.00E-10 | 4.31E-08 |
| GO:0051674 | localization of cell | 365 | -0.51881 | -1.69465 | 1.00E-10 | 4.31E-08 |
| GO:2000145 | regulation of cell motility | 233 | -0.55904 | -1.80180 | 1.00E-10 | 4.31E-08 |
| GO:0006928 | movement of cell or subcellular component | 456 | -0.49124 | -1.61246 | 1.36E-10 | 5.22E-08 |
| GO:0030334 | regulation of cell migration | 222 | -0.56354 | -1.81302 | 1.86E-10 | 6.23E-08 |
| GO:0016477 | cell migration | 336 | -0.51802 | -1.68870 | 1.99E-10 | 6.23E-08 |
| GO:0051270 | regulation of cellular component movement | 254 | -0.53733 | -1.73692 | 4.48E-10 | 1.29E-07 |
| GO:0051493 | regulation of cytoskeleton organization | 176 | -0.57902 | -1.84455 | 5.70E-10 | 1.51E-07 |
| GO:0030036 | actin cytoskeleton organization | 191 | -0.56148 | -1.79487 | 1.88E-09 | 4.63E-07 |
| GO:0030029 | actin filament-based process | 207 | -0.55552 | -1.78211 | 4.22E-09 | 9.71E-07 |
| GO:0030335 | positive regulation of cell migration | 136 | -0.59348 | -1.86614 | 6.09E-09 | 1.28E-06 |
| GO:0040017 | positive regulation of locomotion | 143 | -0.59294 | -1.86954 | 6.34E-09 | 1.28E-06 |
| GO:0032970 | regulation of actin filament-based process | 118 | -0.62183 | -1.93776 | 7.70E-09 | 1.47E-06 |
| GO:0033043 | regulation of organelle organization | 409 | -0.47702 | -1.56211 | 1.27E-08 | 2.31E-06 |
| GO:2000147 | positive regulation of cell motility | 141 | -0.59225 | -1.86585 | 1.35E-08 | 2.33E-06 |
| GO:0007015 | actin filament organization | 127 | -0.59352 | -1.85858 | 1.91E-08 | 3.14E-06 |
| GO:0044087 | regulation of cellular component biogenesis | 313 | -0.49691 | -1.61671 | 2.09E-08 | 3.28E-06 |
| GO:0051272 | positive regulation of cellular component movement | 145 | -0.57704 | -1.82083 | 3.93E-08 | 5.89E-06 |
| GO:0032956 | regulation of actin cytoskeleton organization | 108 | -0.61788 | -1.91404 | 4.14E-08 | 5.94E-06 |
| GO:0051128 | regulation of cellular component organization | 703 | -0.42887 | -1.41855 | 8.23E-08 | 1.14E-05 |
| GO:0019886 | antigen processing and presentation of exogenous peptide antigen via MHC class II | 14 | 0.87773 | 2.59650 | 8.57E-08 | 1.14E-05 |
| GO:0051130 | positive regulation of cellular component organization | 339 | -0.48565 | -1.58351 | 1.40E-07 | 1.79E-05 |
| GO:0007010 | cytoskeleton organization | 381 | -0.46347 | -1.51540 | 4.73E-07 | 5.70E-05 |
| GO:0110053 | regulation of actin filament organization | 91 | -0.62552 | -1.91148 | 4.79E-07 | 5.70E-05 |
| GO:0044089 | positive regulation of cellular component biogenesis | 176 | -0.53450 | -1.70273 | 5.01E-07 | 5.76E-05 |
| GO:0032879 | regulation of localization | 740 | -0.42002 | -1.39041 | 6.43E-07 | 7.15E-05 |
| GO:0002501 | peptide antigen assembly with MHC protein complex | 10 | 0.91701 | 2.44862 | 8.68E-07 | 9.35E-05 |
| GO:0019884 | antigen processing and presentation of exogenous antigen | 24 | 0.74652 | 2.55763 | 1.13E-06 | 1.18E-04 |
| GO:0050900 | leukocyte migration | 102 | -0.58435 | -1.80237 | 1.47E-06 | 1.49E-04 |
| GO:0002504 | antigen processing and presentation of peptide or polysaccharide antigen via MHC class II | 16 | 0.82332 | 2.53292 | 1.58E-06 | 1.56E-04 |
| GO:0002478 | antigen processing and presentation of exogenous peptide antigen | 20 | 0.75876 | 2.48296 | 3.19E-06 | 3.05E-04 |
| GO:0008154 | actin polymerization or depolymerization | 79 | -0.61307 | -1.84898 | 4.00E-06 | 3.73E-04 |
| GO:0022604 | regulation of cell morphogenesis | 86 | -0.60384 | -1.83605 | 4.19E-06 | 3.80E-04 |
| GO:0007166 | cell surface receptor signaling pathway | 656 | -0.41407 | -1.36819 | 4.39E-06 | 3.88E-04 |
| GO:0002495 | antigen processing and presentation of peptide antigen via MHC class II | 15 | 0.82598 | 2.49269 | 6.68E-06 | 5.76E-04 |
| GO:0071674 | mononuclear cell migration | 54 | -0.66636 | -1.93019 | 1.14E-05 | 9.60E-04 |

**Supplementary Table 7**: Characteristics of participants.

| Sample | Cohort | Age at sampling | Sample type sequenced |
|---|---|---|---|
| 1 | EpiHealth | 71-75 | CD4+ T lymphocyte Enriched |
| 2 | EpiHealth | 71-75 | CD4+ T lymphocyte Enriched |
| 3 | EpiHealth | 81-85 | CD4+ T lymphocyte Enriched |
| 4 | EpiHealth | 71-75 | CD4+ T lymphocyte Enriched |
| 5 | EpiHealth | 76-80 | CD4+ T lymphocyte Enriched |
| 6 | EpiHealth | 76-80 | CD4+ T lymphocyte Enriched |
| 7 | EpiHealth | 76-80 | CD4+ T lymphocyte Enriched |
| 8 | EpiHealth | 71-75 | CD4+ T lymphocyte Enriched |
| 9 | EpiHealth | 76-80 | CD4+ T lymphocyte Enriched |
| 10 | EpiHealth | 71-75 | CD4+ T lymphocyte Enriched |
| 11 | EpiHealth | 71-75 | CD4+ T lymphocyte Enriched |
| 12 | EpiHealth | 71-75 | CD4+ T lymphocyte Enriched |
| 13 | EpiHealth | 76-80 | CD4+ T lymphocyte Enriched |
| 14 | EpiHealth | 71-75 | CD4+ T lymphocyte Enriched |
| 15 | EpiHealth | 76-80 | CD4+ T lymphocyte Enriched |
| 16 | EpiHealth | 76-80 | CD4+ T lymphocyte Enriched |
| 17 | EpiHealth | 76-80 | CD4+ T lymphocyte Enriched |
| 18 | EpiHealth | 66-70 | CD4+ T lymphocyte Enriched |
| 19 | EpiHealth | 76-80 | CD4+ T lymphocyte Enriched |
| 20 | EpiHealth | 76-80 | CD4+ T lymphocyte Enriched |
| 21 | EpiHealth | 51-55 | CD4+ T lymphocyte Enriched |
| 22 | EpiHealth | 76-80 | CD4+ T lymphocyte Enriched |
| 23 | EpiHealth | 76-80 | CD4+ T lymphocyte Enriched |
| 24 | EpiHealth | 71-75 | CD4+ T lymphocyte Enriched |
| 25 | EpiHealth | 66-70 | CD4+ T lymphocyte Enriched |
| 26 | EpiHealth | 71-75 | CD4+ T lymphocyte Enriched |
| 27 | EpiHealth | 76-80 | CD4+ T lymphocyte Enriched |
| 28 | EpiHealth | 76-80 | CD4+ T lymphocyte Enriched |
| 29 | EpiHealth | 76-80 | CD4+ T lymphocyte Enriched |
| 30 | EpiHealth | 71-75 | CD4+ T lymphocyte Enriched |
| 31 | UCAN | 76-80 | CD4+ T lymphocyte Enriched |
| 32 | UCAN | 76-80 | CD4+ T lymphocyte Enriched |
| 33 | UAD | 76-80 | PBMCs |
| 34 | UAD | 71-75 | PBMCs |
| 35 | UAD | 66-70 | PBMCs |
| 36 | UAD | 81-85 | PBMCs |
| 37 | UAD | 71-75 | PBMCs |
| 38 | UAD | 61-65 | PBMCs |
| 39 | UAD | 56-60 | PBMCs |
| 40 | UAD | 86-90 | PBMCs |
| 41 | UAD | 81-85 | PBMCs |
| 42 | UAD | 71-75 | PBMCs |
| 43 | UAD | 86-90 | PBMCs |
| 44 | UAD | 71-75 | PBMCs |
| 45 | UAD | 76-80 | PBMCs |
| 46 | UAD | 81-85 | PBMCs |
| 47 | UAD | 81-85 | PBMCs |
| 48 | UAD | 81-85 | PBMCs |
| 49 | UAD | 66-70 | PBMCs |
| 50 | UAD | 81-85 | PBMCs |
| 51 | UAD | 66-70 | PBMCs |
| 52 | UAD | 76-80 | PBMCs |
| 53 | UAD | 76-80 | PBMCs |
| 54 | UAD | 91-95 | PBMCs |
| 55 | UAD | 76-80 | PBMCs |
